# Evaluation of Residual Disinfectant Levels in Public Drinking Water Distribution Systems in Relation to an *Acanthamoeba* Keratitis Outbreak—Illinois 2002–2009

**DOI:** 10.1101/2021.08.27.21262491

**Authors:** Hannah Holsinger, Anna Blackstock, Susan Shaw, Sharon L. Roy

## Abstract

*Acanthamoeba* keratitis (AK) is a painful, potentially blinding eye disease associated with contact lens use and corneal injury. AK, caused by the free-living amoeba *Acanthamoeba*, is ubiquitous in the environment and has been isolated from municipal water supplies. It can be tolerant of normal chlorine levels in drinking water. An AK outbreak investigated in 2003–2005 in five Illinois counties showed a lower AK age-standardized rate ratio in Cook County than in surrounding counties and was hypothesized to be due in part to reductions in residual disinfectant levels (RDLs) in drinking water. We evaluated RDLs in public water systems in the same five Illinois counties over eight years (2002–2009) using a multivariable model of water system RDL measurements. Fitted RDLs for each county were in the acceptable range by United States Environmental Protection Agency standards for the entire study period. After correcting for multiple testing, two of the surrounding counties had fitted RDLs that differed from Cook County for one year—this pattern differed from the epidemiologic pattern of cases observed in the AK outbreak. Our findings do not support the hypothesis that the development of AK was associated with changes in RDLs in the five Illinois counties.

## Introduction

*Acanthamoeba* keratitis (AK) is a rare eye infection that can lead to corneal scarring and loss of sight (Bacton et al.1993). It is caused by the amoeba *Acanthamoeba. Acanthamoeba* is ubiquitous in the environment and can be found in soil (Sawyer 1989), air (Kingston and Warhurst 1969), sewage (Gaze et al. 2011), fresh and brackish water (John and Howard 1995), swimming pools (Cerva 1971), hot tubs (Samples et al. 1984), and tap water (Seal et al. 1999, Shoff et al. 2008, Stockman et al. 2011, Trzyna et al. 2010). *Acanthamoeba* has been detected in both water and biofilms growing on pipe surfaces at various points in drinking water treatment plants and distribution systems (Thomas and Ashbolt 2011) where, in general, the amoebae have been shown to increase in numbers at distal points (Power and Nagy 1999; Långmark et al. 2007).

*Acanthamoeba* can survive a range of harsh conditions including extreme temperatures (Brown and Cursons 1977), salinity (Bergmanson et al. 2011), antimicrobial agents (Ficker et al. 1990), and disinfectants (De Jonckheere and Van De Voorde 1976). The *Acanthamoeba* cyst is much more tolerant than the trophozoite form, but both forms can be tolerant of normal chlorine residual levels found in tap water. Studies have shown that trophozoites are inactivated in free chlorine concentrations of 1.25 mg/L (Cursons et al. 1980) to 1.4 mg/L (Derreumaux et al. 1974) after 30 minutes. However, as part of their normal life cycle and under adverse conditions, trophozoites can encyst. Cysts have been shown to survive up to 75 mg/L of free chlorine for 7 days (Kilvington and Price 1990). The maximum residual disinfectant level (MRDL) of chlorine and chloramine allowed in drinking water is 4 mg/L based on a running annual average of monthly averages of all samples, computed quarterly (USEPA 1998, 2001a). Therefore, cysts are only affected at disinfectant levels much higher than levels found in drinking water distribution systems (Trzyna et al. 2010).

AK has been associated with eye trauma (Bharathi et al 2009) and contact lens use (Thebpatiphat et al. 2007). A multi-state outbreak investigation in the United States found 89% of AK cases involved contact lens users (Verani et al. 2009). AK has been associated with rinsing contact lenses in tap water (Seal et al. 1999), wearing contact lenses while swimming, and use of homemade contact lens solution (Stehr-Green et al. 1987). Many commercial multipurpose contact lens solutions (MPS) have limited disinfection efficacy against *Acanthamoeba* (Johnston et al. 2009, Shoff et al. 2008). A study evaluating AK in non-contact lens users found the main AK risk factors were eye trauma and ocular surface disease (Erdem et al. 2014).

In 2006, an outbreak of AK was detected in Illinois when the Department of Ophthalmology at the University of Illinois at Chicago (UIC) noted a considerable increase in the number of AK cases seen between June 1, 2003, and November 30, 2005 (Joslin et al. 2006). Subsequent investigation revealed it to be a nationwide outbreak (Verani et al. 2009). Cases seemed to increase across the United States in 2003 and 2004, and the MPS Advanced Medical Optics^®^ Complete MoisturePlus™ was shown to be the main risk factor and was voluntarily recalled. Concurrent with the nationwide investigation, the AK outbreak cases in Chicago and surrounding counties were evaluated and the four counties around Chicago were found to have had higher AK age-standardized rate ratios than the referent county (Cook County, which includes Chicago) (Joslin et al. 2006). Since the risk of AK appeared to vary based on geographic location, investigators speculated that water quality might be playing a role and conjectured that the increase in AK, particularly outside of Cook County, might be partially explained by reductions in tap water disinfection and by the effects of long-distance drinking water distribution (Joslin et al. 2006, 2007).

The first two theories involved a temporal association between the increase in cases of AK and the implementation of the United States Environmental Protection Agency’s (EPA) Stage 1 Disinfectant and Disinfection Byproduct Rule (DBPR) that went into effect between 2002 and 2004 (Joslin et al. 2006, 2007, 2010; Shoff et al. 2008; Thebpatiphat et al. 2007). The Stage 1 and 2 DBPR are intended to reduce the public’s exposure to disinfection byproducts. Animal studies have shown disinfection byproducts to be carcinogenic, causing adverse reproductive and developmental effects. The Stage 1 and 2 DBPR established MRDLs for common water disinfectants (USEPA 1998, 2001a, 2006). In order to comply with the Stage 1 DBPR, public water systems have options to reduce the disinfection byproducts by modifying their disinfection practices, including, but not limited to, reducing the contact time or concentration of the disinfectant they are using, or switching to a different type of disinfectant (USEPA 2006). For example, water systems could use chloramines, rather than chlorine, as a secondary disinfectant in water distribution pipelines (Seidel et al. 2005). EPA also requires certain public water systems using surface water to maintain minimum residual disinfectant levels in their water distribution systems. The residual disinfectant level (RDLs) entering the distribution system can be no lower than 0.2 mg/L for more than 4 hours and the level within the distribution system must not be undetectable in more than 5% of water samples within a month for two consecutive months (USEPA 2004). During the study period Illinois had additional requirements for systems chlorinating to have a minimum free chlorine residual of 0.1 mg/L at distant points in the distribution systems, 0.2 mg/L minimum free chlorine residual for chlorinating systems using surface waters, and a minimum free chlorine residual of 0.4 mg/L in storage tanks (IAC 1995). The frequency of residual disinfectant monitoring within each system depends on the size of the population served (USEPA 2001b, 2004). Investigators theorized that differences in the AK age-standardized rate ratios observed in Illinois between Cook County and the four outlying counties might have resulted from changes in the microbial environment, potentially due to either reduced disinfectant levels or the transition from chlorine to chloramine as a secondary disinfectant (Joslin et al. 2006). Another hypothesis was related to the distance between the location of water disinfection and the end user. Investigators reasoned that biofilms were more highly developed in the distal parts of the water distribution system, hypothesizing that the observed geographic variation in AK rates between Cook County and the four outlying counties may be due to the distance between water treatment and use (Joslin et al. 2006).

To explore these three hypotheses, we evaluated RDLs in the public drinking water systems in Cook County and the four surrounding counties to determine if decreases in RDLs occurred over time or between the counties.

## Materials and Methods

RDLs from January 1, 2002 through September 25, 2009 (the entire study period), were obtained from the Illinois Environmental Protection Agency (Illinois EPA) for public drinking water systems within five counties including and surrounding Chicago (Cook, DuPage, Lake, McHenry, and Will). These were the same five counties of residence of the AK case-patients assessed at UIC during June 1, 2003–November 30, 2005 (Joslin et al. 2006). Data provided by Illinois EPA included water system name, county, population served, water source, date of water sample measurement, type of residual disinfectant used (chlorine or chloramine), and free and/or total chlorine levels measured in the distribution system. Total chlorine is the combination of chlorine that is free to disinfect (called free chlorine) plus chlorine combined with ammonia and certain other nitrogenous compounds (called combined chlorine or chloramine) (CDC 2014). Some public water systems were missing data (i.e., either a free or a total chlorine measurement was unavailable or not reported) for 1–3 years during the study period (indicated by the “Neither” category in Figure 1). Since the number of water systems reporting free chlorine was more stable compared to total chlorine over time, free chlorine was used for the analysis and is henceforth referred to as the residual disinfectant level (RDL). Due to variation in the populations served by each of the systems, the number of measurements reported per year by each system also varied—systems serving small populations had few measurements whereas systems serving large populations had many. For each system, the median of the RDLs for each year was used as the outcome in statistical models.

**Figure 1.**
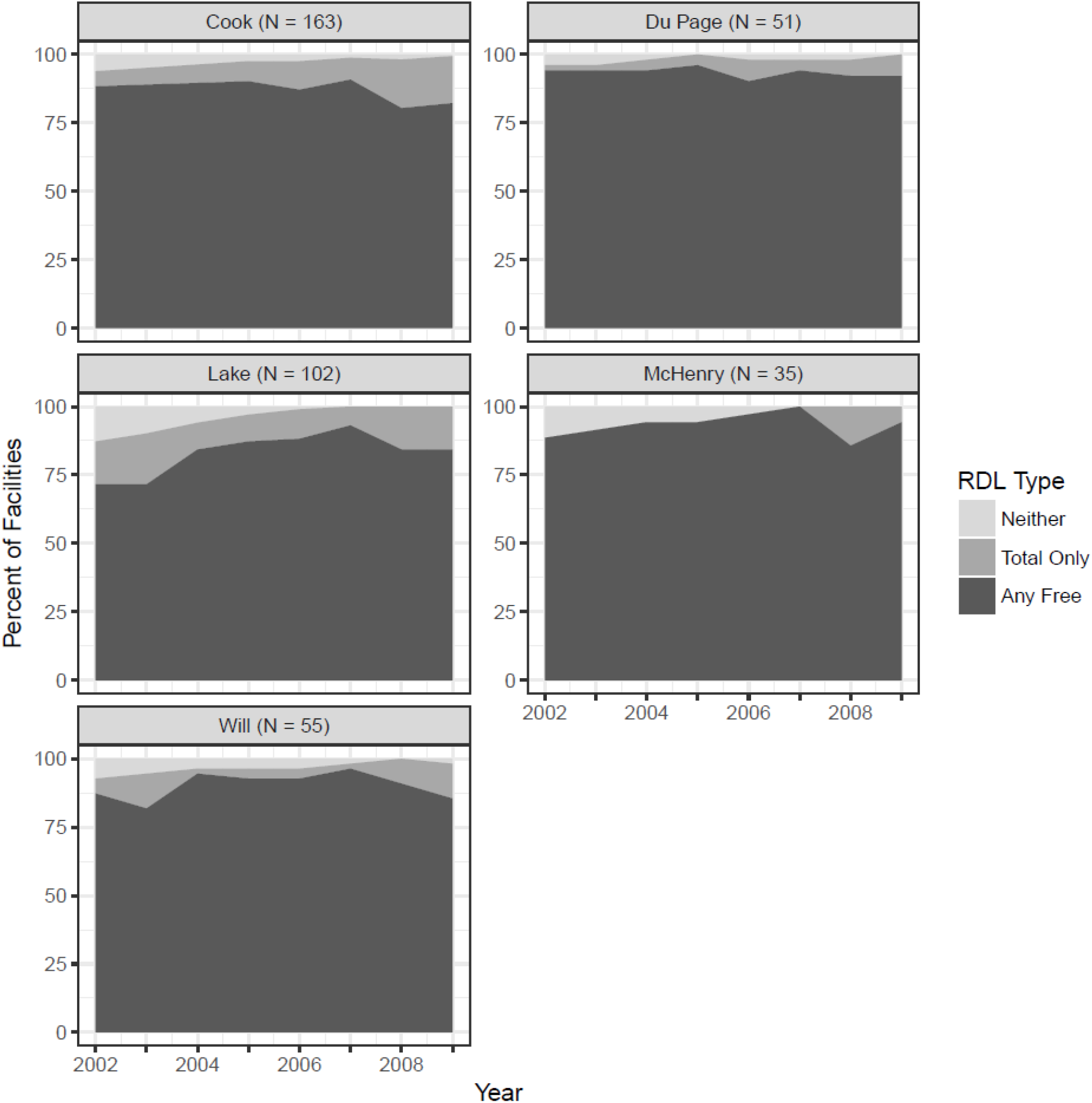
Number of Facilities Reporting Free Chlorine, Total Chlorine, or Neither Residual Disinfectant Levels during the Study Period.

A multivariable model for annual facility RDL measurement was chosen from several candidate models. Each model was fit using generalized estimating equations (GEE) to account for the fact that facilities had repeated measurements over time (Liang and Zeger 1986). Since both location and time were of interest, each model included county and year. Other variables under consideration were number of measurements per facility per year (divided into quartiles), water source (ground or surface water), and two-way interactions involving county, year, and water source. Various correlation structures were also considered. The quasilikelihood information criterion (QIC) was used to choose the final model (Pan 2001), which used an exchangeable correlation structure and included county, year, and the interaction between county and year. Using this model, the fitted mean RDLs of the five counties (using Cook as a reference) were compared for each year individually, and the yearly fitted mean RDLs were compared (using 2002 as a reference) within each county. The False Discovery Rate (FDR) was used to control for multiple comparisons (Benjamini and Hochberg 1995). To assess whether or not the variability within systems would affect the results, resampling was used to fit 1000 models with the same three covariates (county, year, and the county-year interaction), each with a randomly chosen single RDL measurement for each facility for each year of the study.

## Results

Data were available for 434 public water systems in the five counties from January 1, 2002–September 25, 2009. Eight systems were inactive and 20 were missing county information. For the remaining 406 systems, 218 surface water and 188 ground water systems, a total of 532,685 RDLs were collected. Residual disinfection type was available for 325 systems—the majority used chlorine and a small minority used chloramines (around 1.5%). Table 1 shows descriptive statistics for the 406 active systems.

**Table 1.** Public Water System Descriptions and *Acanthamoeba* Keratitis Age-Standardized Rate Ratios, by County—Illinois, 2002–2009.

| County | Cook | DuPage | Lake | McHenry | Will | Total |
| --- | --- | --- | --- | --- | --- | --- |
| # Systems | 163 | 51 | 102 | 35 | 55 | 406* |
| # Surface Water Systems (%) | 139 (85.3) | 36 (70.6) | 36 (35.3) | 0 (0) | 7 (12.7) | 218 (53.7) |
| # Ground Water Systems (%) | 24 (14.7) | 15 (29.4) | 66 (64.7) | 35 (100) | 48 (83.3) | 188 (46.3) |
| Number of AK cases<sup>†</sup> June 03–Nov 05 | 14 | 8 | 2 | 2 | 5 | 31 |
| Age-Standardized Rate Ratios<sup>§</sup> | 1.00 | 3.59 ** | 1.31 | 2.69 | 3.66 ** | - |

\*Out of the 434 public water systems evaluated during the study period, 20 had missing county information and an additional eight were listed as inactive in the Illinois EPA database at the time of analysis. These 28 systems were excluded from the estimated populations in Table 1.
<sup>†</sup>AK: *Acanthamoeba* keratitis. Numbers of cases diagnosed between June 1, 2003 and November 30, 2005 (Joslin 2006).
§ Joslin, C.E., Tu, E.Y., McMahon, T.T., Passaro, D.J., Stayner, L.T., Sugar, J. 2006 Epidemiological characteristics of a Chicago-area *Acanthamoeba* keratitis outbreak. Am J Ophthalmol. 142, 212–217.
\*\* Statistically significant at alpha = 0.05.

During the study period, 76 of the 406 systems (19%) reported 134 samples (134/532,685; 0.03%) with RDLs > 4 mg/L and 43 systems (11%) reported 61 samples (0.01%) with RDLs ≥ 10 mg/L, far above the 4 mg/L MRDL. It is conceivable that RDL levels might be over the acceptable maximum annual average of 4 mg/L, but RDLs of 10 mg/L or more were not expected and are likely due to reporting error and therefore were excluded from the analysis. Conversely, 262 systems (65%) reported 28,504 samples (5%) with RDLs of < 0.2 mg/L (the minimum residual disinfectant limit for certain surface water systems) and 96 systems (24%) reported 1108 samples (0.2%) with RDLs of 0 mg/L. The overall median RDL was 0.63 mg/L, and 99% of values were from 0.03 to 2.1 mg/L.

The model containing year, county, and the county-year interaction was found to be best by QIC and was chosen as the final model. Table 2 shows estimated mean RDLs and confidence intervals resulting from the final model for each county during all years evaluated, and Figure 2 displays these values graphically. While Lake, McHenry, and Will Counties had at least one year with fitted mean RDLs significantly different than those of Cook County for the same year before correcting for multiple comparisons, only Lake County in 2004 had an estimated mean RDL that was significantly different and lower than Cook County’s value after correcting for multiple comparisons using an FDR threshold of 0.05. McHenry County in 2009 was also significantly different from Cook County after FDR correction, but McHenry County values were higher than Cook County in all years. Comparing years within counties yielded several significant results. All counties except for Cook County had at least one year with fitted mean RDLs significantly different than those of 2002 within the same county. However, after FDR correction only two counties, McHenry and Will, had fitted mean RDLs significantly different than those in 2002. McHenry County had significantly higher fitted mean RDL values in 2005, 2007, and 2009 than in 2002, and Will County had fitted mean values from 2004–2009 that were all significantly lower than the 2002 measurement and that all remained significantly lower after FDR correction. The plots of estimates and confidence intervals based on resampling models revealed similar relationships between counties and over time.

**Table 2.**
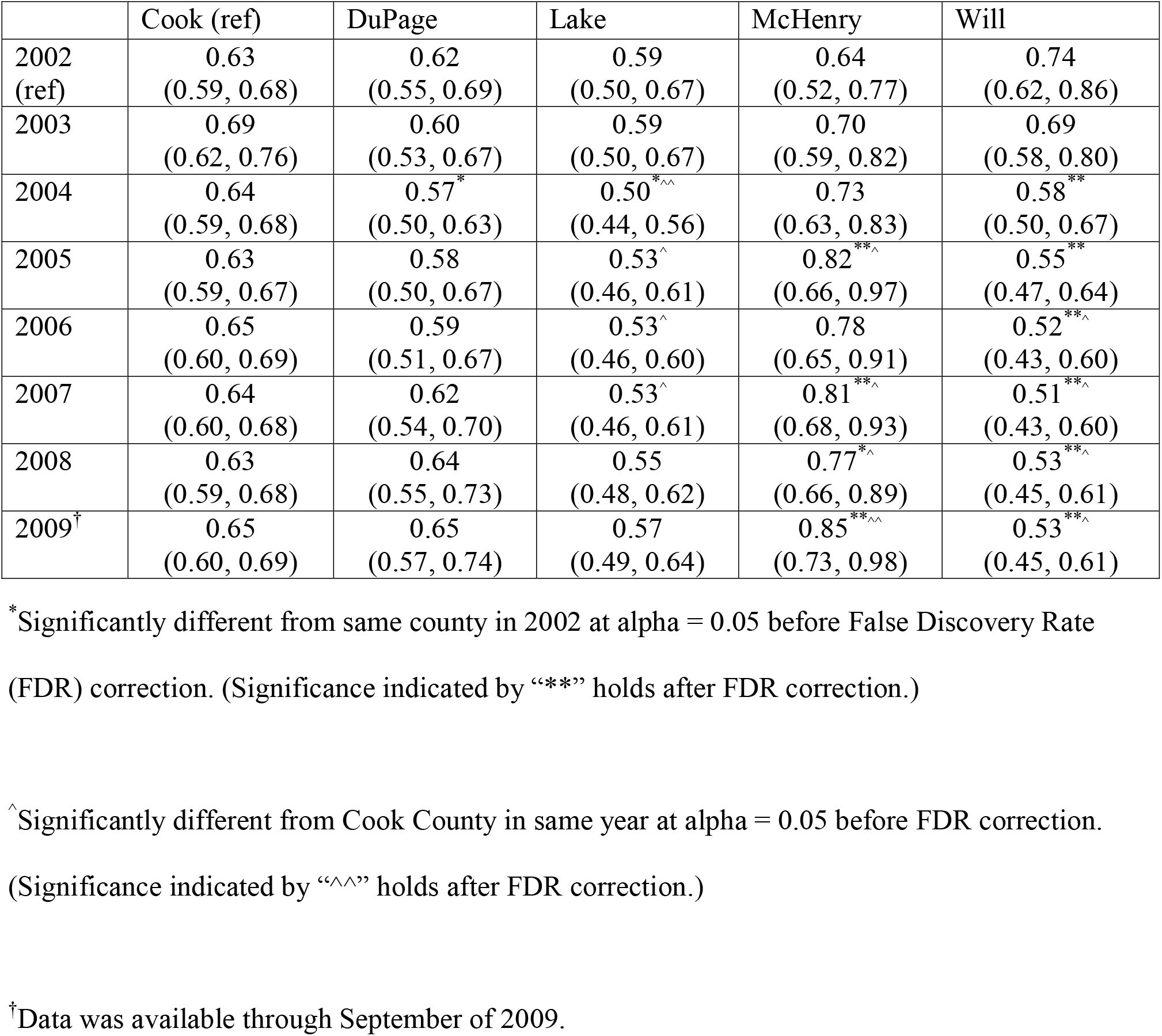
Estimated Mean Residual Disinfectant Levels and 95% Confidence Intervals from Public Water Systems in Five Counties—Illinois, 2002 – 2009. Table 2 shows estimated mean residual disinfectant levels (RDLs) (mg/L) and their confidence intervals. The estimated means were obtained from the model containing county, year, and the county by year interaction.

**Figure 2.**
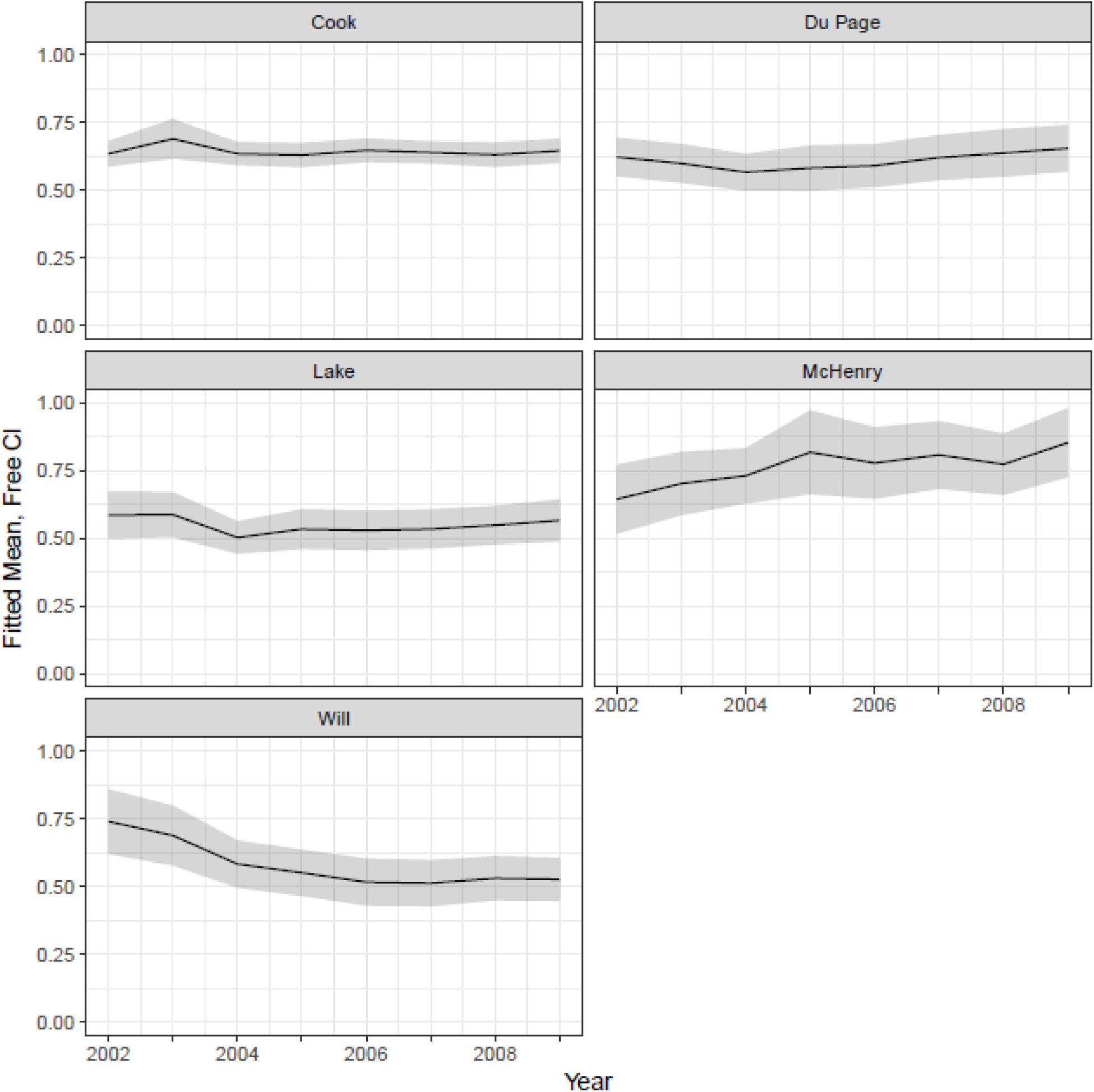
Estimated Mean Residual Disinfectant Levels for Five Counties—Illinois, 2002 – 2009^*^. ^*^Data was available through September of 2009.

## Discussion

The aim of this study was to evaluate RDLs in public drinking water systems in the five counties including and surrounding Chicago to determine if significant reductions had occurred in these levels over time or between the counties that could explain the observed geographic discrepancy in the AK age-standardized rate ratios. UIC investigators put forth several water-related hypotheses for why AK rate ratios were higher in the outlying counties compared to Cook County. We focused on three of these hypotheses—decreased RDLs, a change in the disinfectant type, and increased distance between water treatment and use (Joslin et al. 2006).

Results of our fitted model did not indicate a dramatic shift in RDLs during 2002 through September 2009, nor was there a pattern of reduction in RDLs that mirrored the pattern of increased AK age-standardized rate ratios observed in the outlying counties during the Illinois outbreak investigation as displayed in Figure 2. After FDR correction for multiple testing, only one county had a significantly lower RDL than Cook County in only one year during 2002–2009. Although McHenry and Will Counties each had at least one year with a fitted mean annual RDL significantly different from that of 2002 after FDR correction, the pattern was inconsistent with and therefore not supportive of the working hypothesis that reductions in RDLs were associated with AK risk. In Will County, the fitted mean annual RDLs were significantly lower in 2004– 2009, but in McHenry County, the RDLs were higher in 2005, 2007, and 2009.

Another hypothesis put forward to explain the epidemiology of AK observed in Illinois during 2003–2005 was a switch in secondary disinfectant type. The majority of all public water systems evaluated used chlorine as a secondary disinfectant. The City of Chicago Department of Water Management provided water to the majority of the public water systems. Only McHenry County, a ground water system, did not receive any water from the City of Chicago or Lake Michigan. The City of Chicago Department of Water Management has treated its water with chlorine for >30 years (Verani et al. 2009) and made no change in disinfection chemicals (Methvin 2009). Therefore, the type of chemical disinfectant used by the majority of the 406 public water systems evaluated in the five-county area remained unchanged during the entire study period, during periods of changing regulations associated with the Stage 1 DBPR that went into effect during 2002–2004 (U.S. EPA 1998, 2001a, 2006).

The third hypothesis concerning the risk factors for AK in Illinois involved distance from water treatment and the risk for *Acanthamoeba*-contaminated water. The UIC investigators hypothesized that the outlying counties, being a greater distance from Chicago, had greater distances between water treatment and the end users in their respective counties, thereby contributing to the observed increased risk for AK compared to the referent Cook County (Joslin et al. 2006). Loss of residual disinfectant and microbial regrowth in the drinking water distribution system has many causes including water age (LeChevallier 2003; NRC 2005) and distance from treatment (Långmark et al. 2007; Power and Nagy 1999). This hypothesis could not be directly evaluated with the data collected for this evaluation because county is not a proxy for distance from treatment. Although most water was initially treated by the City of Chicago Department of Water Management, booster residual disinfectant stations are used in some systems in Illinois to maintain RDLs and not every county received water from the City of Chicago Department of Water Management (IEPA 2015). Without knowing which water system served each AK-case patient and where each person resided, evaluations of issues possibly contributing to RDL degradation and biofilm development in each distribution system or premise plumbing serving those patients are not possible.

Not knowing which water systems served the AK-case patients was the most problematic limitation of this study. Private water systems were also not included, and it is possible that some of the AK-case patients identified by UIC (Joslin et al. 2006) were private well owners and not represented in our dataset. This was an ecological study, and interpretation of the results was hindered by the limitation of all such studies, namely the fallacy that group characteristics are shared by individuals. Although ecologic studies cannot demonstrate a causal relationship, they can identify promising avenues of research that might further our understanding of causal relationships. However, our study found no evidence of systematic reductions in RDLs and does not support the hypothesis that reductions in RDLs that were hypothesized to be associated with Stage 1 DBPR regulations led to the AK outbreak. Another limitation is the number and frequency of RDL measurements taken varied depending on system size. Therefore, the measurements analyzed in this study over-represent large public water systems. Using median RDLs helped to control for systematic bias that may have been introduced into the calculation had mean RDLs been used.

Although no apparent pattern that would support the hypothesized relationship between reduced RDLs and the development of AK was identified, there is no question that *Acanthamoeba* spp. are present in some water systems and that tap water could be one of the sources of exposure. Several studies have isolated *Acanthamoeba* in water distribution systems (Seal et al. 1992; Shoff et al. 2008; Stockman et al. 2011; Trzyna et al. 2010). Free-living amoebae, including *Acanthamoeba* spp., appear to be common in premise plumbing. *Acanthamoeba* can provide protection and aid in life cycles for amoeba-resistant microorganisms such as *Legionella* (Pruden et al. 2013). Further research is needed to understand the role of biofilm growth and *Acanthamoeba* keratitis.

Although tap water is a possible source of exposure to *Acanthamoeba*, (Cerva 1971; Gaze et al. 2011; John and Howard 1995; Kingston and Warhurst 1969; Samples et al. 1984; Sawyer 1989), the high level of chlorine tolerance (De Jonckheere and Van De Voorde 1976) likely limits the feasibility of mitigating *Acanthamoeba* colonization in water distribution systems, given current chemical disinfection practices. Since *Acanthamoeba* are unlikely to be eliminated from the environment, other methods to limit eye exposure to water should be prioritized. Established guidelines should be followed by contact lens users to help reduce the risk of eye infections.

Good lens-handling practices are important because the efficacy of contact lens disinfection solutions against *Acanthamoeba* has been questioned. Two studies investigating the nationwide outbreak of AK (Joslin et al. 2007; Verani et al. 2009) both found a particular brand of MPS to be the primary cause of the outbreak. Studies of a variety of brands of MPS (Cursons et al. 1980; Shoff et al. 2008) showed that these solutions in general are ineffective disinfectants of *Acanthamoeba*. No standard protocols for efficacy testing of contact lens solutions against *Acanthamoeba* currently exist. Furthermore, neither the United States Food and Drug Administration nor the International Organization for Standardization requires testing of the efficacies of contact lens solutions against *Acanthamoeba* species (Cursons et al. 1980; Shoff et al. 2008). Standardized procedures for determining the disinfection efficacy of contact lens solutions against *Acanthamoeba* would likely improve this situation for public health. Following the voluntary recall in May 2007 of the brand of MPS implicated in the AK outbreak, CDC worked with a group of reference laboratories and ophthalmologic institutions to monitor the occurrence of laboratory-confirmed AK at these sites. From these data, it appeared that the collective number of AK cases at these sites failed to return to pre-outbreak levels following the product recall (Yoder et al. 2012). There could have been many reasons for this, including surveillance artifact, but the general ineffectiveness of MPSs as disinfectants of *Acanthamoeba* was hypothesized to play a primary role (Johnston et al. 2009). The failure of a main barrier against *Acanthamoeba* is of great concern, particularly since it remains ubiquitous in the environment. Prevention of future cases of AK requires contact lens solutions that are effective disinfectants of *Acanthamoeba* coupled with continued emphasis on proper lens care practices and hygiene.

Our ecological study found no evidence of systematic reductions in RDLs in the outbreak areas and does not support the hypothesis that there were reductions in RDLs resulting from actions taken by public water systems to comply with the EPA’s Stage 1 Disinfectant and Disinfection Byproduct Rule or because of changes in disinfection chemical type that might have affected the microbial environment in the distribution systems (Joslin et al. 2006).

Nevertheless, *Acanthamoeba* spp. are known contaminants of drinking water distribution systems regardless of geography (Seal et al. 1992; Shoff et al. 2008, Stockman et al. 2011, Trzyna et al. 2010) and both trophozoite and cyst forms are known to be tolerant of chlorine at regulated levels within distribution systems. Therefore, tap water mitigation is not a practical or timely way to reduce *Acanthamoeba* exposure. Other strategies to reduce exposure and prevent *Acanthamoeba* amplification should be prioritized, including reinforcing appropriate contact lens hygiene, removing contact lenses before contact with water, and developing MPSs that are more efficacious at disinfecting *Acanthamoeba*. Guidelines for contact lens wearers to prevent AK and other eye infections are available at http://www.cdc.gov/contactlenses/protect-your-eyes.html.

## Data Availability

The data used in this manuscript were obtained from the state of Illinois via EPA's Region 5 office.

## Acknowledgements

We wish to acknowledge data collection conducted by Illinois Environmental Protection Agency, as well as Janet Kuefler from U.S. EPA Region 5.

## Disclaimer

The views expressed in this paper are those of the individual authors and do not necessarily reflect the views and policies of the U.S. Environmental Protection Agency or the Centers for Disease Control and Prevention. This work was partially developed under Cooperative Agreement No. X3 83388101 awarded by the U.S. Environmental Protection Agency to the Association of Schools of Public Health.

